# Tissue-Specific Failure Phenotypes of the Knee Extensor Mechanism Across Skeletal Maturity in Anterior Knee Pain

**DOI:** 10.64898/2026.02.03.26345528

**Authors:** Shinsuke Sakoda, Hiroto Kumagae, Kimiaki Kawano

## Abstract

**Background:** Anterior knee pain (AKP) in young athletes comprises heterogeneous conditions arising within the knee extensor mechanism (KEM). Although skeletal maturity influences AKP prevalence, its relationship with tissue-specific clinical phenotypes remains unclear.

**Methods:** A total of 1,589 patients presenting with sports-related knee injuries were analyzed. AKP was classified into bony and non-bony phenotypes according to the anatomical site of maximal tenderness, used as a clinical surrogate for the primary symptomatic tissue within the KEM. Multivariable logistic regression analyses evaluated associations of skeletal maturity, age, sex, and sports participation type with overall and phenotype-specific AKP.

**Results:** AKP was diagnosed in 577 patients (36.3%), including 274 with bony AKP and 242 with non-bony AKP. Skeletal immaturity was independently associated with overall AKP (adjusted odds ratio [OR], 2.77; p < 0.001). Phenotype-stratified analyses demonstrated a markedly stronger association between skeletal immaturity and bony AKP (adjusted OR, 5.97; p < 0.001), whereas no significant association was observed with non-bony AKP. Participation in repetitive high-energy deceleration sports showed contrasting associations, with a positive association with bony AKP (adjusted OR, 1.55; p = 0.034) and an inverse association with non-bony AKP (adjusted OR, 0.60; p < 0.001).

**Conclusion:** Tissue-dominant AKP phenotypes within the KEM differ according to skeletal maturity. The developmental association of AKP is primarily driven by a predominance of osseous-dominant phenotypes, whereas soft-tissue–dominant phenotypes persist across maturation stages. These findings indicate that developmental differences in AKP reflect variations in tissue-specific clinical vulnerability within a shared mechanical system.

## 1. Introduction

Anterior knee pain (AKP) is one of the most prevalent sports-related conditions in adolescent athletes and represents a major cause of functional limitation, performance decline, and training restriction. ^1, 2^ Despite its high clinical burden, AKP remains difficult to stratify because it encompasses a wide spectrum of conditions traditionally classified as separate diagnostic entities, including traction apophysitis, patellar tendinopathy, and nonspecific AKP. ^3–5^

Conventional diagnostic frameworks primarily categorize AKP according to anatomical labels or imaging findings. ^6, 7^ However, such classifications may obscure shared biomechanical mechanisms because many AKP-related disorders arise within a common load-transmitting structure: the knee extensor mechanism (KEM). ^8–10^ The KEM functions as an integrated kinetic chain that repeatedly transmits tensile forces generated during running, jumping, and deceleration, thereby creating a unified mechanical environment across various athletic activities. ^9–12^

Although mechanical loading conditions are broadly similar across growth stages, the clinical manifestations of AKP vary markedly with skeletal maturation. ^10, 11^ Traction-related osseous disorders such as Osgood–Schlatter disease and Sinding-Larsen– Johansson disease predominantly affect skeletally immature athletes, whereas tendon-dominant pathologies become more common after physeal closure. ^13–16^ This developmental heterogeneity cannot be fully explained by differences in activity level alone and suggests that tissue-specific vulnerability within the KEM may vary across maturation stages. ^9, 10, 12^

We hypothesized that AKP should be conceptualized as a set of distinct structural phenotypes arising from a shared mechanical environment, in which the primary site of mechanical failure shifts developmentally according to tissue vulnerability. Under this framework, osseous structures represent preferential failure sites during skeletal immaturity, whereas soft tissues become relatively more vulnerable following maturation.

To test this hypothesis, we analyzed a large clinical cohort of patients with sports-related knee injuries and classified AKP according to the anatomical site of maximal tenderness as a clinically practical surrogate for the primary pain-generating tissue. Using phenotype-stratified logistic regression analyses, we investigated whether skeletal maturity and sports-related mechanical loading were differentially associated with bony and non-bony AKP phenotypes.

## 2. Methods

### 2.1. Study Design and Participants

This single-center retrospective observational study included consecutive patients who presented to our institution with sports-related knee injuries between January 2017 and November 2025. A total of 1,589 cases were eligible for analysis. Patients were included if demographic data, clinical diagnoses, physical examination findings, and radiographic evaluations were available. Patients with tumors, infections, or a history of prior knee surgery were excluded.

The study protocol was approved by the institutional review board of our institution and conducted in accordance with the Declaration of Helsinki. Informed consent was obtained using an opt-out approach.

### 2.2. Definition of AKP

AKP was defined as self-reported pain localized to the anterior aspect of the knee that was provoked or exacerbated by sports activities or related movements. Imaging abnormalities were not required for diagnosis. AKP was diagnosed clinically after excluding acute traumatic structural injuries requiring alternative diagnoses. ^3–6^

### 2.3. Assessment of Skeletal Maturity

Skeletal maturity was determined by the status of the proximal tibial physis on plain radiographs at initial evaluation, and participants were categorized as having open or closed physes. ^14–16^

### 2.4. Classification of AKP Based on Maximal Tenderness

Patients diagnosed with AKP were further classified according to the site of maximal tenderness identified during standardized physical examination. ^17, 18^ Bony AKP was defined as localized tenderness involving osseous components of the KEM, including the patellar bony region (inferior pole or bipartite patella) and the proximal tibia, particularly the tibial tuberosity. ^13–16^

Non-bony AKP was defined as localized tenderness of the soft tissue structures of the knee extensor mechanism, including the patellar tendon and anterior periarticular tissues, without definite osseous involvement. ^3, 4, 11, 12^

When multiple tender sites were identified, the primary pain generator was determined based on symptom reproducibility during functional activities. ^17, 18^ Patients with AKP in whom no localized tenderness could be identified were excluded from phenotype-specific analyses.

### 2.5. Definition of Sports Type

Participation in repetitive high-energy deceleration (RHD) sports was defined as engagement in sports characterized by frequent jumping, rapid deceleration, cutting, pivoting, and high-intensity directional changes that impose substantial repetitive mechanical loading on the KEM. ^9, 10^ Representative sports included basketball, volleyball, handball, soccer, rugby, badminton, tennis, and gymnastics. Sports not meeting these criteria were classified as non-RHD sports; representative examples included baseball, track and field events, and swimming.

### 2.6. Outcome Definitions

To examine phenotype-specific associations, three outcome variables were defined: overall AKP, defined as the presence of anterior knee pain; bony AKP, defined as osseous-dominant AKP; and non-bony AKP, defined as soft tissue–dominant AKP.

### 2.7. Statistical Analysis

Categorical variables were compared using chi-square tests. Logistic regression analyses were performed to identify factors associated with AKP using the three predefined outcome definitions. Both univariate and multivariable logistic regression models were constructed. Multivariable models were adjusted for age, sex, physeal status, and sports type (repetitive high-energy deceleration sports). Odds ratios (ORs) with 95% confidence intervals (CIs) were calculated. A p value < 0.05 was considered statistically significant.

All statistical analyses were performed using R (version 4.3.1; R Foundation for Statistical Computing, Vienna, Austria). A two-sided p-value < 0.05 was considered statistically significant.

## 3. Results

### 3.1. Participant Characteristics

A total of 1,589 patients with sports-related knee injuries were included in the analysis. The mean age was 15.4 ± 3.6 years, and 1,164 patients were male. Open physes were observed in 713 patients. Participation in repetitive high-energy deceleration sports was reported in 1,093 patients.

Among the study population, 577 patients (36.3%) were diagnosed with anterior knee pain (AKP). Based on tenderness classification, 274 patients were categorized as having bony AKP and 242 as having non-bony AKP. Sixty-one patients with AKP in whom no localized tenderness was identified were excluded from phenotype-specific analyses.

Baseline demographic and clinical characteristics are summarized in Table 1.

**Table 1.** Baseline demographic and clinical characteristics of patients with sports-related knee injuries included in the study.

| Variable | Value |
| --- | --- |
| Age, years (mean ± SD) | 15.4 ± 3.6 |
| Male sex (n, %) | 1,164 (73.3) |
| Open physes (n, %) | 713 (44.9) |
| RHD sports (n, %) | 1,093 (68.8) |
| AKP positive (n, %) | 577 (36.3) |
| Bony AKP (n, %) | 274 (47.5) |
| Non-bony AKP (n, %) | 242 (41.9) |
| No tenderness (n, %) | 61 (10.6) |

Percentages for AKP subtypes are calculated among patients diagnosed with AKP.
Abbreviations: AKP, anterior knee pain; RHD, repetitive high-energy deceleration.

### 3.2. Factors Associated With Overall AKP

In univariate logistic regression analyses, younger age, male sex, skeletal immaturity, and sports type were significantly associated with overall AKP.

In multivariable logistic regression analysis adjusting for age, sex, physeal status, and sports type, skeletal immaturity remained independently associated with AKP (adjusted OR, 2.77 95% CI 1.85–4.15, p < 0.001). Younger age also independently predicted AKP (adjusted OR 0.85 per year increase, p < 0.001). Participation in RHD sports was independently associated with a lower prevalence of AKP (adjusted OR 0.64 p < 0.001), whereas sex was not independently associated after adjustment (p = 0.175).

Detailed results are presented in Table 2A.

**Table 2.** Results of multivariable logistic regression analyses examining factors associated with (A) overall anterior knee pain (AKP), (B) bony AKP, and (C) non-bony AKP.

| Variable | Adjusted OR | 95% CI | P value |
| --- | --- | --- | --- |
| Age (per year increase) | 0.85 | 0.80–0.90 | < 0.001 |
| Male sex (vs female) | 1.21 | 0.92–1.60 | 0.175 |
| Open Physes (vs closed) | 2.77 | 1.85–4.15 | < 0.001 |
| RHD sports (vs others) | 0.64 | 0.50–0.82 | < 0.001 |

(B) Bony AKP
| Variable | Adjusted OR | 95% CI | P value |
| --- | --- | --- | --- |
| Age (per year increase) | 0.89 | 0.79–0.99 | 0.038 |
| Male sex (vs female) | 1.46 | 0.89–2.41 | 0.136 |
| Open Physes (vs closed) | 5.97 | 2.80–12.75 | < 0.001 |
| RHD sports (vs others) | 1.55 | 1.03–2.32 | 0.034 |

(C) Non-bony AKP
| Variable | Adjusted OR | 95% CI | P value |
| --- | --- | --- | --- |
| Age (per year increase) | 0.91 | 0.85–0.98 | 0.011 |
| Male sex (vs female) | 1.02 | 0.73–1.43 | 0.900 |
| Open Physes (vs closed) | 0.83 | 0.50–1.37 | 0.462 |
| RHD sports (vs others) | 0.60 | 0.45–0.80 | < 0.001 |

Odds ratios (ORs) with 95% confidence intervals (CIs) were calculated using logistic regression models adjusted for age, sex, physal status, and participation in repetitive high-energy deceleration (RHD) sports.
Closed physes and female sex were used as reference categories.
A two-sided p value < 0.05 was considered statistically significant.

### 3.3. Factors Associated With Bony AKP

In univariate logistic regression analyses, younger age, male sex, and skeletal immaturity were significantly associated with bony AKP, whereas participation in RHD sports showed an inverse association.

When the outcome was restricted to bony AKP, skeletal immaturity demonstrated a markedly stronger independent association (adjusted OR 5.97, 95% CI 2.80–12.75, p < 0.001). Participation in RHD sports was also independently associated with an increased prevalence of bony AKP (adjusted OR 1.55, p = 0.034). Younger age remained significantly associated (adjusted OR 0.89, p = 0.038), whereas sex showed no independent association.

Detailed results are presented in Table 2B.

### 3.4. Factors Associated With Non-Bony AKP

In univariate logistic regression analyses, younger age was significantly associated with non-bony AKP, whereas skeletal maturity and sex showed no meaningful associations. Participation in RHD sports demonstrated an inverse association.

In multivariable analysis adjusting for age, sex, physeal status, and sports type, skeletal maturity showed no independent association with non-bony AKP (adjusted OR 0.83, p = 0.462). Participation in RHD sports remained inversely associated (adjusted OR 0.60, p < 0.001). Younger age independently predicted non-bony AKP (adjusted OR 0.91, p = 0.011), whereas sex was not independently associated.

Detailed results are presented in Table 2C.

### 3.5. Phenotype-Specific Patterns of Association

Comparative analyses revealed distinct phenotype-dependent patterns. Skeletal immaturity showed a strong independent association with bony AKP but not with non-bony AKP. Conversely, RHD sports were positively associated with bony AKP but inversely associated with non-bony AKP (Figure 1).

**Figure 1.**
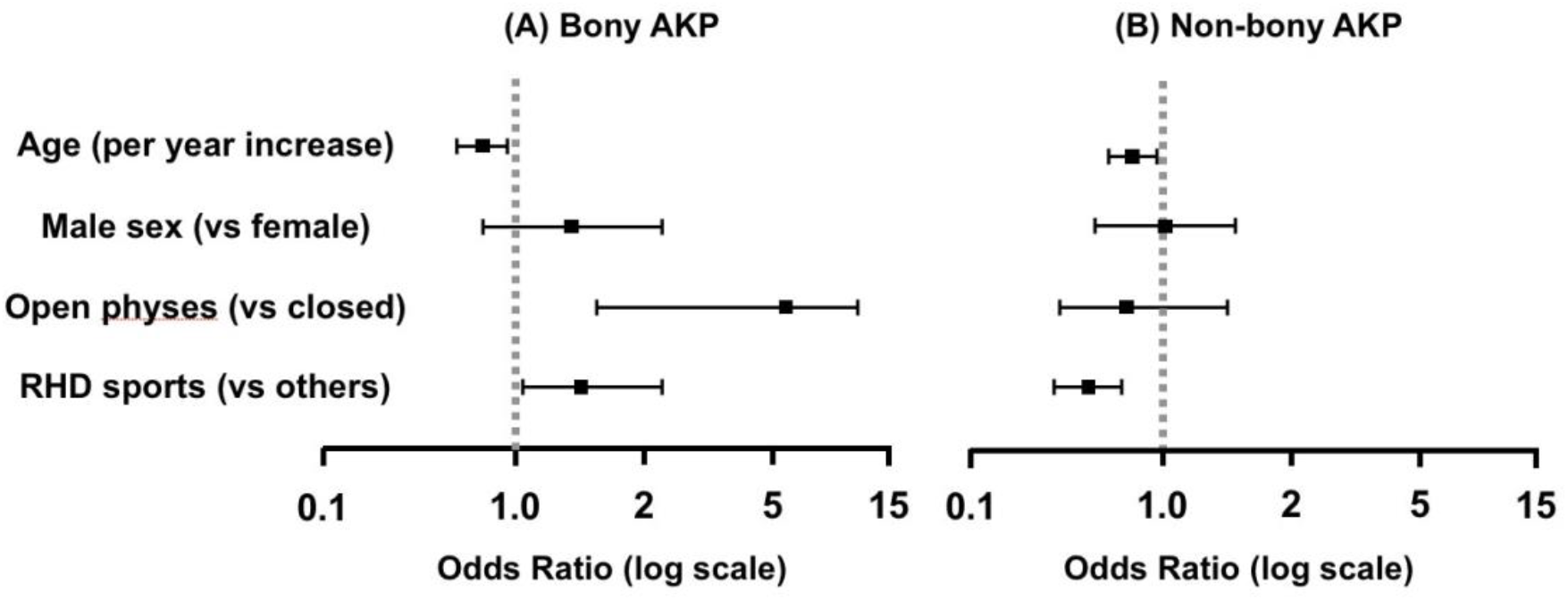
Multivariable logistic regression analysis of factors associated with phenotype-specific anterior knee pain Forest plots showing adjusted odds ratios (ORs) with 95% confidence intervals (CIs) for factors associated with (A) bony anterior knee pain (AKP) and (B) non-bony AKP. The vertical dashed line indicates the reference value (OR = 1.0). Odds ratios greater than 1 indicate a positive association with the outcome, whereas values less than 1 indicate a negative association. All models were adjusted for age, sex, physeal status, and participation in repetitive high-energy deceleration (RHD) sports. ORs are presented on a logarithmic scale.

**Figure 2.**
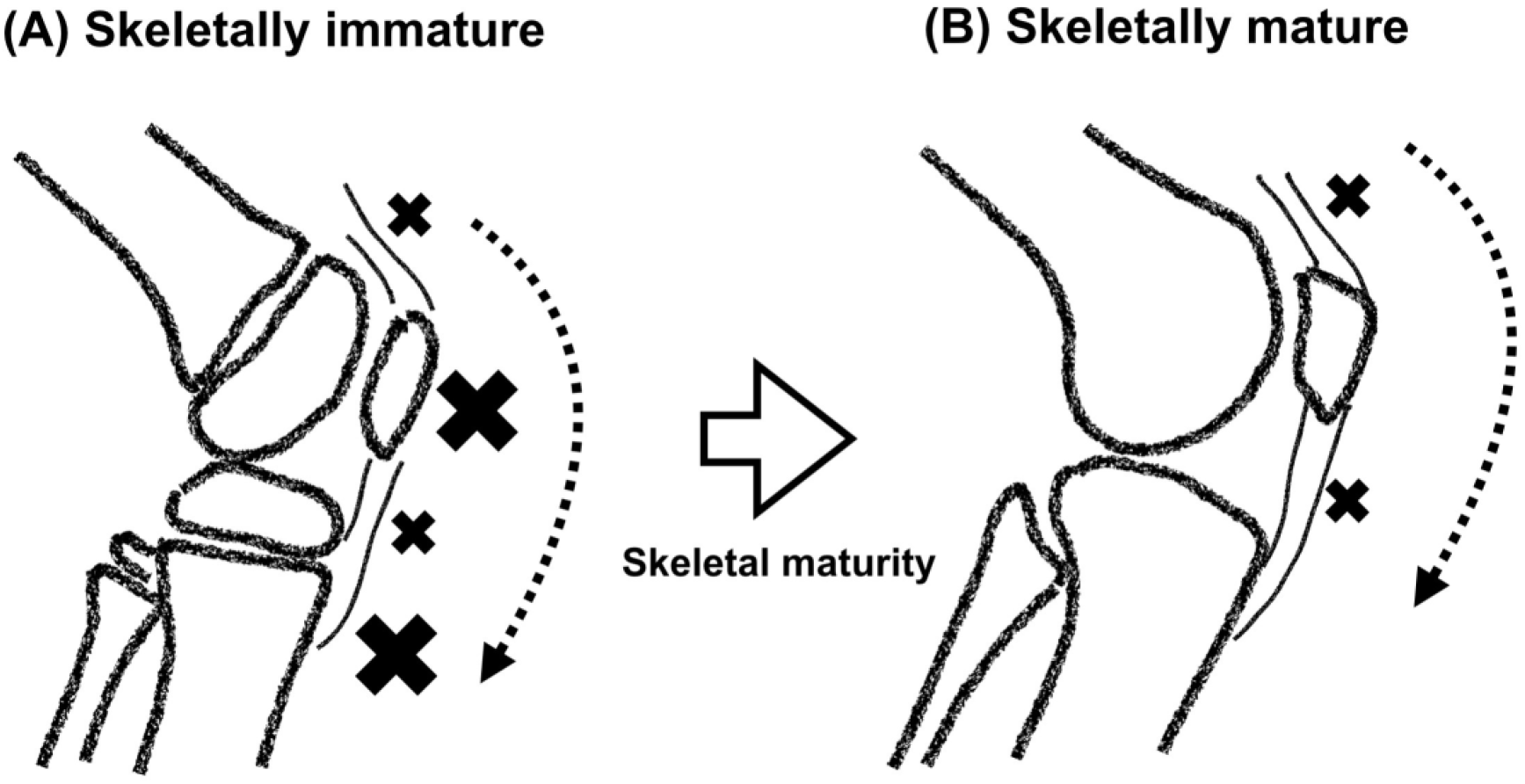
Schematic summary of phenotype-specific tissue involvement across skeletal maturity (A) In skeletally immature athletes, clinical phenotypes were more frequently associated with osseous components of the knee extensor mechanism (KEM), particularly apophyseal and tendon–bone insertion regions. (B) In skeletally mature athletes, osseous-dominant phenotypes were less common, whereas soft-tissue–dominant phenotypes remained prevalent. Dashed arrows indicate the anatomical load-transmission pathway of the KEM. Cross marks indicate representative locations of predominant symptomatic sites observed in this study.

## 4. Discussion

### 4.1. Phenotype-Specific Heterogeneity of Anterior Knee Pain

The present study demonstrates that AKP is not a uniform clinical entity but comprises tissue-specific clinical phenotypes that vary according to skeletal maturity. When AKP was stratified by tenderness-based tissue classification, skeletal immaturity showed a strong independent association with bony AKP but no meaningful association with non-bony AKP. These findings indicate that the developmental association of AKP is primarily attributable to osseous-dominant presentations, whereas soft-tissue–dominant presentations remain prevalent across maturation stages.

This divergence suggests that age-related differences in AKP cannot be explained solely by variations in overall mechanical loading. ^9^ If loading magnitude were the principal determinant, similar developmental associations would be expected for both phenotypes. ^8–10^ Instead, the findings indicate that tissue-specific susceptibility during growth influences the clinical manifestation patterns observed within a shared mechanical environment. ^12–15^

### 4.2. Structural Interpretation Within the Knee Extensor Mechanism

The KEM functions as an integrated load-transmitting structure that is repeatedly exposed to substantial tensile forces during athletic activities. ^8–10^ Within this unified mechanical system, clinical manifestations appear to preferentially involve tissues with relatively lower structural tolerance. ^9, 10^

The present findings suggest that, during skeletal immaturity, osseous structures linked to the KEM are more frequently involved in clinical symptom presentation. ^13–15^ Apophyseal cartilage, developing ossification centers, and tendon insertion sites may exhibit reduced tolerance to repetitive traction stress during growth, which may contribute to the higher prevalence of osseous-dominant phenotypes observed in skeletally immature athletes. ^13, 14^

With skeletal maturation, progressive mineralization and structural reinforcement of osseous components may reduce the frequency of bone-dominant clinical presentations. ^14, 15^ In contrast, soft-tissue–dominant presentations remain prevalent across maturation stages, resulting in differences in the relative predominance of tissue-specific phenotypes rather than a replacement of one phenotype by another. ^3, 4, 11^

These observations indicate that developmental differences in structural tolerance are reflected in the anatomical distribution of clinical phenotypes within the KEM. This interpretation provides a clinically coherent framework for understanding phenotype-specific patterns of AKP without assuming direct alterations in biomechanical load pathways.

### 4.3. Orthopedic Integration of Traction-Related Disorders

The present structural perspective facilitates integration of traction-related disorders traditionally considered distinct diagnostic entities. Conditions such as Osgood–Schlatter disease, Sinding-Larsen–Johansson disease, and symptomatic bipartite patella share a common mechanical background characterized by repetitive traction stress acting on developmentally susceptible osseous structures. ^13, 14^

Rather than representing unrelated diseases, these conditions may be interpreted as osseous-dominant clinical phenotypes occurring within a shared mechanical environment during growth. As skeletal maturation progresses and osseous tolerance improves, the frequency of such presentations declines, whereas soft-tissue–dominant phenotypes remain clinically prevalent. ^3, 14, 15^

This integrated interpretation clarifies why clinically distinct diagnostic labels emerge under similar athletic exposures and emphasizes the importance of developmental tissue characteristics in shaping phenotype distribution.

### 4.4. Biomechanical Considerations

From a biomechanical perspective, the KEM distributes tensile forces across interconnected anatomical components during athletic movements. ^8, 9^ The present findings do not suggest that fundamental loading pathways change with maturation. Instead, they indicate that developmental differences in tissue tolerance are reflected in variations in the observed distribution of tissue-specific clinical phenotypes. ^9, 10^

Osseous structures appear more susceptible to symptomatic involvement during skeletal immaturity, whereas soft-tissue–dominant presentations continue to occur after maturation. Thus, age-related differences in AKP should be interpreted primarily as differences in tissue-specific clinical susceptibility rather than as evidence of mechanical redistribution within the extensor mechanism.

### 4.5. Methodological Significance of Tenderness Localization

Tenderness localization should be regarded as a clinically meaningful indicator rather than merely a subjective examination finding. ^17, 18^ In overuse-related AKP, imaging frequently fails to demonstrate definitive structural abnormalities, making physical examination essential for identifying symptomatic tissues. ^3, 6^

Focal tenderness is widely used in routine orthopedic practice to localize pathological structures and is generally reproducible when standardized examination procedures are applied. ^17, 18^ Within the KEM, the site of maximal tenderness may serve as a practical surrogate marker reflecting tissue-specific clinical susceptibility. ^8, 9^

Accordingly, tenderness-based classification offers clinically relevant structural information and enables practical stratification of tissue-dominant AKP phenotypes.

### 4.6. Clinical Implications for Growth-Stage–Specific Load Management

The developmental differences in tissue-specific phenotype distribution have important implications for clinical management. Rather than focusing solely on lesion-specific diagnoses, treatment strategies should prioritize regulation of mechanical loading within the KEM while considering growth-stage–specific tissue susceptibility.

In skeletally immature athletes, the predominance of osseous-dominant presentations suggests that excessive traction stress on developing bone should be mitigated through activity modification, progressive training adjustment, and movement optimization aimed at reducing knee extension loads. ^13–15^

After skeletal maturation, although osseous-dominant presentations become less frequent, soft-tissue–dominant phenotypes remain prevalent. ^3, 4, 11^ Consequently, clinical management should continue to emphasize load regulation and biomechanical optimization rather than isolated treatment of specific tissues. ^8, 9^

Recognition of tissue-specific phenotype patterns may therefore support developmentally appropriate load management strategies and improve clinical stratification of patients with AKP.

### 4.7. Limitations and Future Directions

Several limitations should be acknowledged. Tenderness-based classification represents a clinically inferred surrogate and was not directly validated against imaging or histopathological findings. In cases with multisite tenderness, classification based on a dominant site may not fully capture underlying pathophysiological complexity.

Radiographic physeal status also does not completely represent biological maturation of apophyseal cartilage. ^14^

Future investigations integrating advanced imaging modalities, biomechanical assessments, and longitudinal outcome analyses are warranted to further clarify the structural correlates and prognostic significance of tissue-specific clinical phenotypes.

## 5. Conclusion

Anterior knee pain (AKP) comprises tissue-specific clinical phenotypes whose relative predominance differs according to skeletal maturity. The strong association between skeletal immaturity and AKP is primarily attributable to osseous-dominant phenotypes, whereas soft-tissue–dominant phenotypes remain prevalent across maturation stages.

These findings support interpretation of developmental differences in AKP as variations in tissue-specific clinical susceptibility within a shared mechanical system rather than as evidence of fundamental changes in load-transfer pathways. Recognition of phenotype patterns may facilitate developmentally appropriate load management and improve clinical stratification.

## Data Availability

The data that support the findings of this study are available from the corresponding author upon reasonable request. The dataset contains clinical information derived from a single-center sports injury database and is not publicly available due to institutional privacy regulations and ethical restrictions.

## Research Data Availability

After publication, the datasets generated and analyzed during the current study will be made available from an appropriate public repository in accordance with journal policies.

## CRediT Authorship Contribution Statement

**Shinsuke Sakoda:** Conceptualization, Methodology, Investigation, Formal analysis, Data curation, Visualization, Writing – original draft, Writing – review & editing. **Hiroto Kumagae:** Methodology, Investigation, Data curation – review & editing. **Kimiaki Kawano:** Supervision, Validation – review & editing, Project administration.

## Acknowledgements

The authors thank the clinical staff of Ashiya Central Hospital for their assistance with data collection and patient management. We also appreciate the support provided by the radiology department for imaging assessments.

